# Assessment of Magnitude of Transfusion Transmissible Infections and Associated Factors among Blood Donors at Woliso Blood Bank, South-West Shewa Zone, Oromia, Ethiopia: Across-Sectional Study

**DOI:** 10.1101/2024.02.08.24302506

**Authors:** Alemnesh W/Amlak, Habtamu Oljira, Belay Tafa, Samuel Demissie Darcho, Sisay Dabi Begna

## Abstract

**Background:** Blood transfusion is an effective treatment for saving millions of lives, even though transfusion-transmissible infections are the major problem. The prevalence of transfusion-transmissible infections varies across different geographical populations. This study aims to assess the prevalence of transfusion-transmissible infections and associated factors among blood donors at Woliso Blood Bank, South-West Shewa Zone, Oromia, Ethiopia.

**Method:** An institutional-based cross-sectional study design was conducted. A structured and pretested questionnaire was used to collect data through a face-to-face interview. Data was entered into Epi Data version 3.1, and it was exported to STATA version 17.0 for data cleaning and analysis. A binary logistic regression analysis was carried out to identify factors associated with outcome variables. Accordingly, variables that fulfilled p-values <0.25 on the bivariate logistic regression were considered candidates for multivariate logistic regression to control for possible confounders. The odds ratios along with the 95% confidence interval were used to present the finding, and statistical significance was reported at a p-value of 0.05.

**Results:** The overall prevalence of transfusion-transmissible infections was 9.5% (95% CI: 6.3– 12.9%). Participants with no formal education [AOR=4.84; 95% CI= 1.09, 21.46], unprotected sexual intercourse with multiple partners [AOR=4.77; 95% CI= 1.38, 16.44], and participants with less frequency of blood donation [AOR=2.85; 95% CI: 1.16, 6.99] were significantly associated with transfusion transmissible infections.

**Conclusion:** The prevalence of transfusion-transmissible infections was high in this study area. Educational level, unprotected sexual intercourse with more partners, and a number of blood donations were found to be independent predictors of transfusion-transmitted infections. Blood banks and regional health bureaus should work on community mobilization and enhancing health promotion through prevention and control by considering the identified associated factors.

## Introduction

Transfusions of blood are a vital component of medical care that saves lives and enhances the quality of life for millions of people worldwide. Blood transfusions are most commonly used to treat patients who have had trauma, emergencies, disasters, or accidents; children with severe anemia from malaria or malnutrition; and women who are bleeding during pregnancy or childbirth. It is also used to support complicated medical and surgical operations like transplants and cardiovascular surgery in nations with contemporary healthcare systems. Adequate health care requires timely and universal access to safe blood and blood products, as well as the appropriate use of these resources (1).

Our society places a high priority on transfusion safety, and clinical trials are being conducted to find ways to reduce the risk of known and emerging illnesses in all blood products. Using pathogen reduction methods, the risk of infection from bacterial and viral pathogens can be effectively reduced (2). Blood grouping and compatibility testing, safe blood collection procedures, accurate testing for transfusion-transmissible infections, recruiting and retention of low-risk blood donors, and appropriate use and safe administration of blood are all factors that contribute to blood safety (3).

A 100% voluntary, unpaid blood donation program requires blood donors to be recruited through efficient procedures, community education, communication tactics, and effective research. Additional key elements are standardized procedures for donor selection, quality of blood collection, counselling donors to ensure their health and safety, encourage repeat donations, donor follow-up, and retention strategies to maintain a stable base of safe voluntary blood donors who give blood regularly (4).

In Ethiopia, there are currently 43 blood banks, one central national blood bank (NBB), and forty-two operational regional blood banks, with at least one in each of the country’s administrative regions so that Woliso blood bank is one of the 10 BB in Oromia region (5).

A number of serious public health issues still affect blood donors, including syphilis, hepatitis B, hepatitis C, and HIV. The World Health Organization (WHO) states that screening for syphilis, HBV, HCV, and HIV should be done on all blood donors. The frequency of TTIs has significantly decreased as a result of the WHO’s introduction of stringent donor screening protocols for blood-borne diseases (6).

Blood shortages and contaminated blood are the two most important problems associated with blood transfusions in the developing world, especially in Africa. These problems all too often result in severe health consequences like postpartum hemorrhage deaths or the spread of potentially fatal infections like HIV and hepatitis. It would be possible to avoid these damaging effects on health by taking steps to increase blood availability and safety (7). HIV, HBV, and HCV are very concerning due to their prolonged viremia and carrier or latent states. Additionally, they may result in fatal, persistent, or life-threatening diseases. Because infectious indicators are so common among blood donors in developing nations, blood safety remains a serious issue (8).

The absence of procedures and infrastructure for ensuring a safe blood supply increases the danger of infection spread by transfusion in many countries. These problems include a shortage of knowledgeable staff, inconsistent test kit supplies or the use of low-quality test kits, an unstable supply chain, and inadequate cold chain facilities. A fragmented blood supply system, with varied technical standards and no central monitoring, can further jeopardize safety measures (4).

The World Health Organization (WHO) has established a goal of enhancing regional blood safety by 2012 by strengthening organization and management, blood donor recruitment and collection, donor blood testing, and appropriate clinical blood usage (9).Hepatitis viruses infect approximately 2.3 billion individuals worldwide, resulting in roughly 1.4 million fatalities, 90% of which are caused by hepatitis B and C viruses. As a result, chronic hepatitis infections have raised mortality rates and represent a significant burden on healthcare systems in many countries, including Ethiopia. HBV infects one-third of the world’s population (more than two billion people), posing a severe health threat (10).

The World Health Organization (WHO) announced a policy in 2016 to eliminate viral hepatitis by 2030. Africa, particularly Sub-Saharan Africa, bears a large share of the global burden of viral hepatitis, particularly chronic hepatitis B and C virus infections. Major developments in the management of hepatitis C have put elimination within reach, but several difficulties will need to be navigated on the path to elimination. Many of the challenges faced are unique to sub-Saharan Africa and the development of strategies is complicated by a scarcity of good data from countries and regions within sub-Saharan Africa(11).

In developed countries, prevention of TTIs has been achieved by reducing unnecessary transfusions, using only regularly-screened volunteer donors, excluding donors with risk factors and screening of all donated blood for infection. In many developing nations, however, none of these measures are implemented consistently, and the risk of TTIs remains high (12). At the national blood bank in Ethiopia, the prevalence of the major TTIs (Hepatitis B Virus=5.23 %, HIV=2.29 %t, and Hepatitis C Virus=2.30 %) is high. As a result, regular monitoring of the amount of transfusion transmissible infections in blood donors is critical to prevent infectious disease transmission(7).

The prevalence of infection or the proportion of blood donations with a positive result is directly related to the safety of the blood supply because this has an impact on the residual risk of blood products used for patient care and also on the risk due to errors in blood quarantine and release (even though test-positive donations are discarded). So that the prevalence of an infection in blood donations is dependent on the prevalence of the infection in the population from which donors are selected and on the effectiveness of donor motivation, mobilization and selection processes(13).

Transfusion transmittable infections among blood donors are the subject of limited epidemiological investigations in Ethiopia (14). Majority of the studies from Ethiopia focused on secondary data analysis /trend analysis of TTIs among voluntary blood donors, which may have a limitation to identify associated risk factors. No research was done at Woliso Blood Bank to determine the prevalence and associated factors. Therefore, the aim of this study is to assess prevalence and associated factors of transfusion transmittable infections among blood donors at Woliso Blood Bank of South West Shewa Zone, Oromia Region, Ethiopia, 2021.

## Materials and Methods

### Study setting, design, and period

An institutional-based cross-sectional study was conducted at Woliso Blood Bank in Southwest Shewa. It is one of the 20 zones in Oromia Regional State, located 114 kilometers from Addis Ababa, the country’s capital city. Woliso Blood Bank is a government organization that was founded in 2013 by the Oromia regional health bureau in partnership with the FMoH to promote voluntary, non-remunerated blood donation and blood collection. It aims to serve all populations of 5,561,976 (S/W/Shewa 1,110,112, N/Shewa 1,450,525, W/Shewa 2,381,072, and Orom/F/esp./zone 620,267) by preventing morbidity and death from blood shortages by supplying safe and adequate blood on a timely and free basis depending on hospital demand. Currently, 14 hospitals get blood from the blood bank on a regular basis (5). The study was conducted from November 1-30, 2021.

### Source of Population and Study Population

All volunteer blood donors in Woliso town and catchment district were the source population, and all volunteer blood donors whose age was between 18 and 65 years were the study population.

### Inclusion criteria and Exclusion criteria

Blood donors with an age of greater than or equal to 18 and less than 65 years with good health were included in the study. Additionally, donors after three months with a weight greater than 48 kg and volunteer blood donors who signed consent were included in the study, and participants who were not willing to participate in the study during data collection and those with nausea, vomiting, headache, and discomfort were excluded from the study.

### Sample Size Determination

The required sample size (n) for objective one was calculated using a single population proportion formula with a 95% CI, 3% margin of error, and prevalence of similar studies (12).

n = p (1-p) / d2

n = 3.8416*0.0676/0.0009 = 288

Taking 10% non-response, the final sample size was 0.1*288 + 288 = **317**.

The sample size of the second objective was smaller than the first objective. Therefore, the final sample size by taking 10% non-response was **317.**

### Sampling Procedure/Technique

A systematic sampling technique was used among volunteer blood donors in the blood bank who met the facility’s donation criteria. Using the blood donor database, a sampling frame was built. The blood bank receives an average of 900 blood donations every month. After calculating the k value, a random beginning point was chosen using the lottery method. The following formula is used to determine the sampling interval: K=N/n, where N is the population size (total numbers of collections in the blood bank during data collection time, which is 900), n is the calculated sample size (317), and k is 3. Using this method, each individual in the population had a known and equal chance of being chosen.

### Data Collection Method

#### Data Collectors and Supervisors

Based on their experience with data collection and supervision, three health professional data collectors and one supervisor were assigned.

#### Data collection instruments

The structured questionnaire was adapted from studies conducted before this study (12,16,20) and modified in context. The questionnaire was prepared in English, translated into Afan Oromo for data collection, and then re-translated back to English to check its consistency. It consists of two sections comprising information on the socio-demographic characteristics of the study participants and their exposure to various risk factors.

### Study Variables

#### Dependent variable

Transfusion transmissible infections

#### Independent variables

- Sociodemographic characteristics include age, sex, marital status, education, and occupation; behavioral factors include multiple sexual partners, therapeutic drug injection, alcohol use, cigarette smoking, and chat chewing; clinical factors include previous transfusion, surgery, and health service-related factors; previous donation and post-donation counseling; previous exposure to a sharp injury (cut); history of tooth extraction; and history of blood contact.

### Operational definitions

**Voluntary non-remunerated blood donors** were blood donors who gave blood voluntarily without any payment and not for their own family.

**Replacement donors:** donors of blood who replace used blood by their relatives or friends from blood bank stocks

**Donor screening criteria:** Physical and clinical assessment criteria used to accept or reject a blood donor

**Transfusion-transmitted infection**: blood-borne infections that may be transmitted during the blood transfusion process (human immunodeficiency virus, hepatitis B virus, hepatitis C virus, and syphilis).

**Positive for TTIS**: If the donor samples contain one or more TTIs (the donor sample should be positive for TTIS two times by ELISA test).

### Data quality control

To assure the quality of the data, a one-day training was given to supervisors and data collectors by the principal investigator on the data collection tool. Pre-testing was conducted outside the study area on 5% of the sample size before data collection in the same area of the study, and some amendments were made based on the pre-test findings. The questionnaire was checked daily for accuracy, consistency, and completeness by the supervisor. Furthermore, the supervisor gave feedback and corrections regarding the collected data on a daily basis to the data collectors. Standard operating procedures were strictly followed for TTI screening. For quality assurance, all TTI-positive samples and 10% of the negative samples were collected to be blindly re-examined by other laboratory technologists.

### Data Processing and Analysis

Data was entered into Epi Data version 3.1, and it was exported to STATA version 17.0 for data cleaning and analysis. The study participant’s socio-demographic characteristics were described by descriptive statistics, and the results were presented using a frequency table. A bivariable logistic regression analysis was used to examine the determinants of TTI, and a p-value below 0.25 was entered into the multivariable logistic regression model. Multivariable logistic regression analysis was used to examine the association between the determinant variables of TTI. A p-value of less than 0.05 in multivariable logistic regression analysis was considered statistical significance. Both crude and adjusted odds ratios were presented with a 95% confidence interval. Hosmer Leme show goodness-of-fit was used to test for model fitness.

### Ethics approval and consent to participate

The ethical approval and clearance were obtained from the Ambo University College of Medicine and Health Science research and ethical review committee. In addition to that, informed, voluntary, written, and signed consent was obtained from each study participant after a brief explanation of the objective of the study, which assures that participation was voluntary. The participant was informed that questionnaires were confidential and that they had the right to refuse to respond to the questionnaire or participate in the study at any time they wanted. Post-donation counseling was given to those positive donors in a confidential manner. Finally, the test result was kept in soft copy using Pass Word as well as in hard copy by locking it at Woliso Blood Laboratory.

## Results

### Socio-demographic factors

Three hundred seventeen voluntary blood donors participated in this study from 317 sampled participants, with a 100 percent response rate. The study consisted of 230 (72.6%) males and 87 (27.4%) females. The mean (± standard deviation) age of the participants was 28.40 (± 9.34) years. The majority of study participants (98.1%) were urban dwellers and around two-fifths (46.1%) were students. Around half (48.3%) of the participants donated blood for the first time (**Table 1**)

**Table 1.**
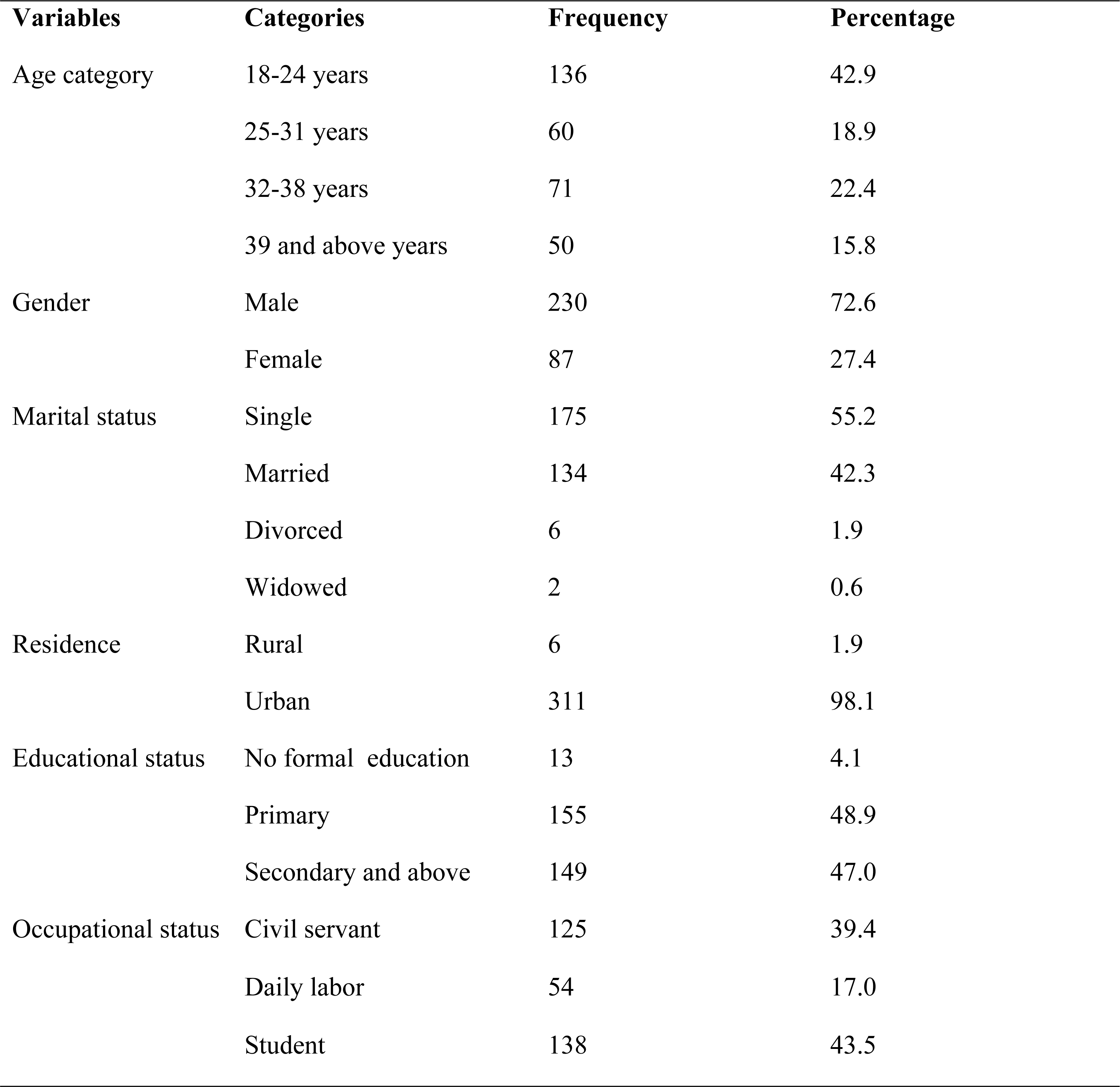
Socio-demographic characteristics of voluntary blood donors at Woliso Blood Bank of South West Shewa, Oromia Region, Ethiopia, 2021.

| Variables | Categories | Frequency | Percentage |
| --- | --- | --- | --- |
| Age category | 18-24 years | 136 | 42.9 |
|  | 25-31 years | 60 | 18.9 |
|  | 32-38 years | 71 | 22.4 |
|  | 39 and above years | 50 | 15.8 |
| Gender | Male | 230 | 72.6 |
|  | Female | 87 | 27.4 |
| Marital status | Single | 175 | 55.2 |
|  | Married | 134 | 42.3 |
|  | Divorced | 6 | 1.9 |
|  | Widowed | 2 | 0.6 |
| Residence | Rural | 6 | 1.9 |
|  | Urban | 311 | 98.1 |
| Educational status | No formal education | 13 | 4.1 |
|  | Primary | 155 | 48.9 |
|  | Secondary and above | 149 | 47.0 |
| Occupational status | Civil servant | 125 | 39.4 |
|  | Daily labor | 54 | 17.0 |
|  | Student | 138 | 43.5 |

### Clinical and behavioural characteristics of study participants

Among the study participants, around 4.1% had received blood products, and around 1.9% were exposed to unsafe injections. Tooth extraction, post-donation counseling, had sex, history of STD in family, and unprotected sex habit were found in 10.4%, 21.8%, 53.3%, 10.4%, and 14.8% of the study participants, respectively. (**Table 2**)

**Table 2.**
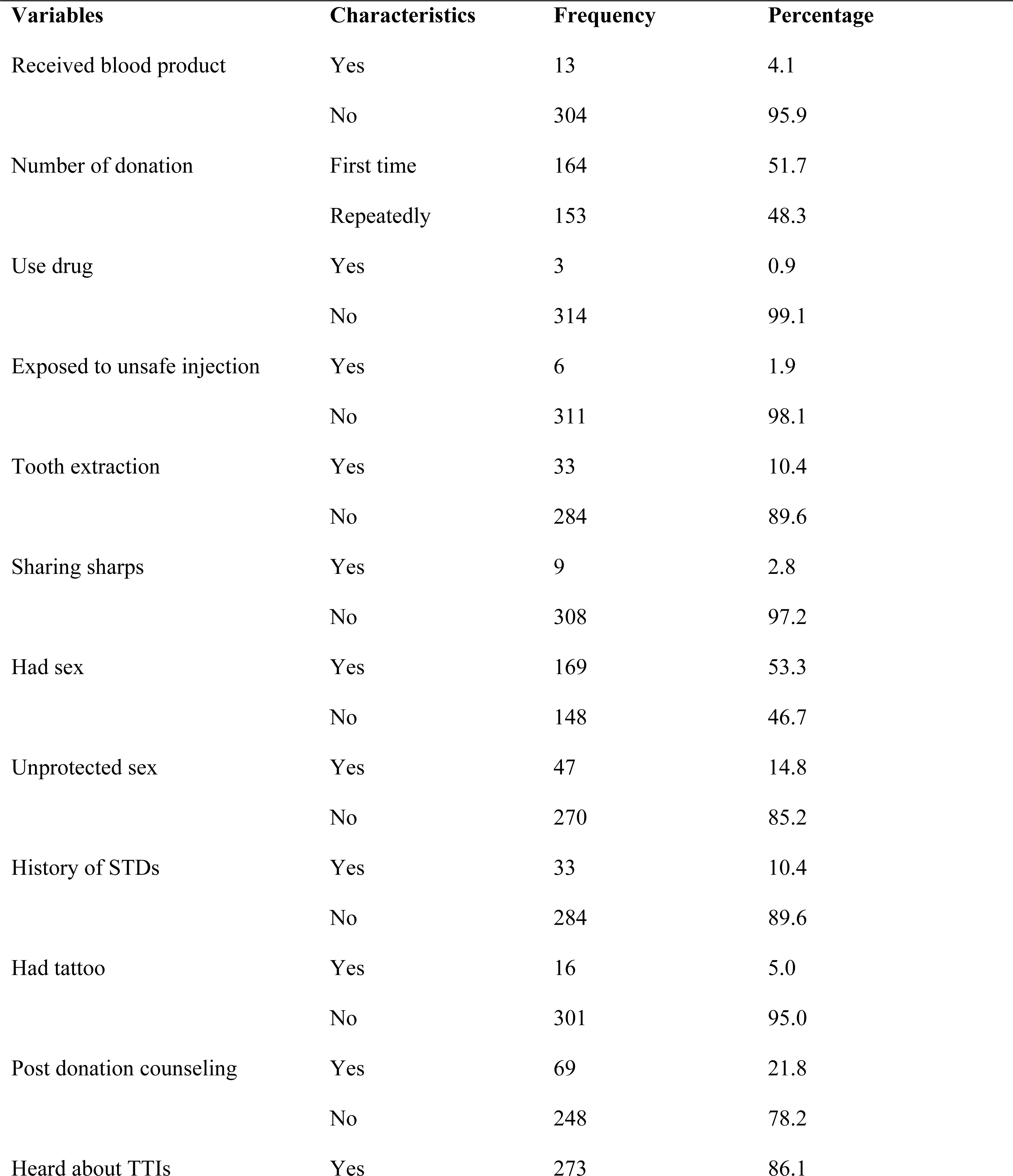
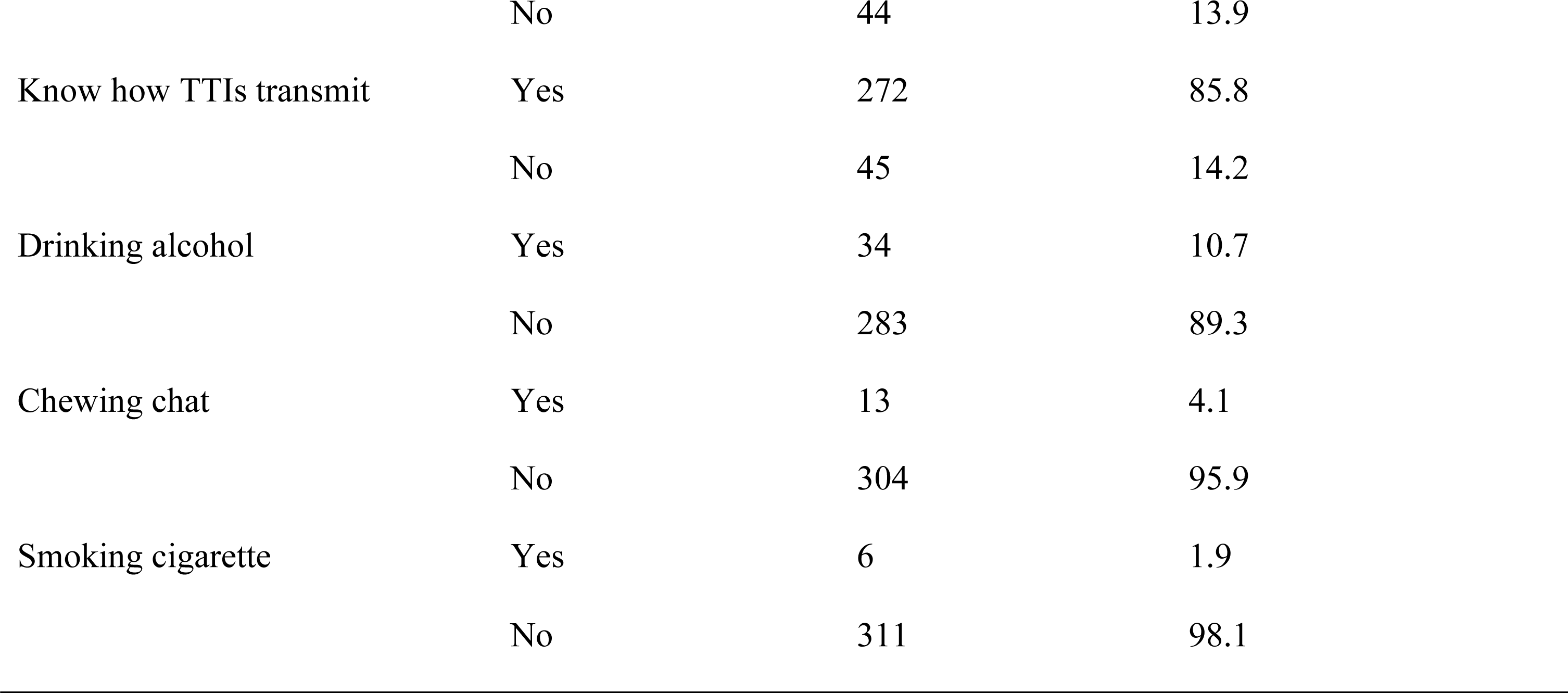
Clinical history and behavioral characteristics of voluntary blood donors at Woliso Blood Bank of South West Shewa, Oromia Region, Ethiopia, 2021.

| Variables | Characteristics | Frequency | Percentage |
| --- | --- | --- | --- |
| Received blood product | Yes | 13 | 4.1 |
|  | No | 304 | 95.9 |
| Number of donation | First time | 164 | 51.7 |
|  | Repeatedly | 153 | 48.3 |
| Use drug | Yes | 3 | 0.9 |
|  | No | 314 | 99.1 |
| Exposed to unsafe injection | Yes | 6 | 1.9 |
|  | No | 311 | 98.1 |
| Tooth extraction | Yes | 33 | 10.4 |
|  | No | 284 | 89.6 |
| Sharing sharps | Yes | 9 | 2.8 |
|  | No | 308 | 97.2 |
| Had sex | Yes | 169 | 53.3 |
|  | No | 148 | 46.7 |
| Unprotected sex | Yes | 47 | 14.8 |
|  | No | 270 | 85.2 |
| History of STDs | Yes | 33 | 10.4 |
|  | No | 284 | 89.6 |
| Had tattoo | Yes | 16 | 5.0 |
|  | No | 301 | 95.0 |
| Post donation counseling | Yes | 69 | 21.8 |
|  | No | 248 | 78.2 |
| Heard about TTIs | Yes | 273 | 86.1 |
|  | No | 44 | 13.9 |
| Know how TTIs transmit | Yes | 272 | 85.8 |
|  | No | 45 | 14.2 |
| Drinking alcohol | Yes | 34 | 10.7 |
|  | No | 283 | 89.3 |
| Chewing chat | Yes | 13 | 4.1 |
|  | No | 304 | 95.9 |
| Smoking cigarette | Yes | 6 | 1.9 |
|  | No | 311 | 98.1 |

### Sero-prevalence of HIV, HBV, HCV and syphilis among voluntary blood donors

The overall sero-prevalence of transfusion-transmissible infections (the proportion of voluntary blood donors with at least one transfusion-transmissible infection marker) was 9.5% (95% CI: 6.3, 12.9). The prevalence of HBV, syphilis, HIV, and HCV was 5.05% (95% CI: 3.0, 7.0), 3.15% (95% CI: 1.0, 5.0), 0.63% (95% CI: 0.0, 2.0), and 0.63% (95% CI: 0.0, 2.0), respectively. In this study, co-infections were not detected in all study participants. (**Fig 1**)

**Figure 1.**
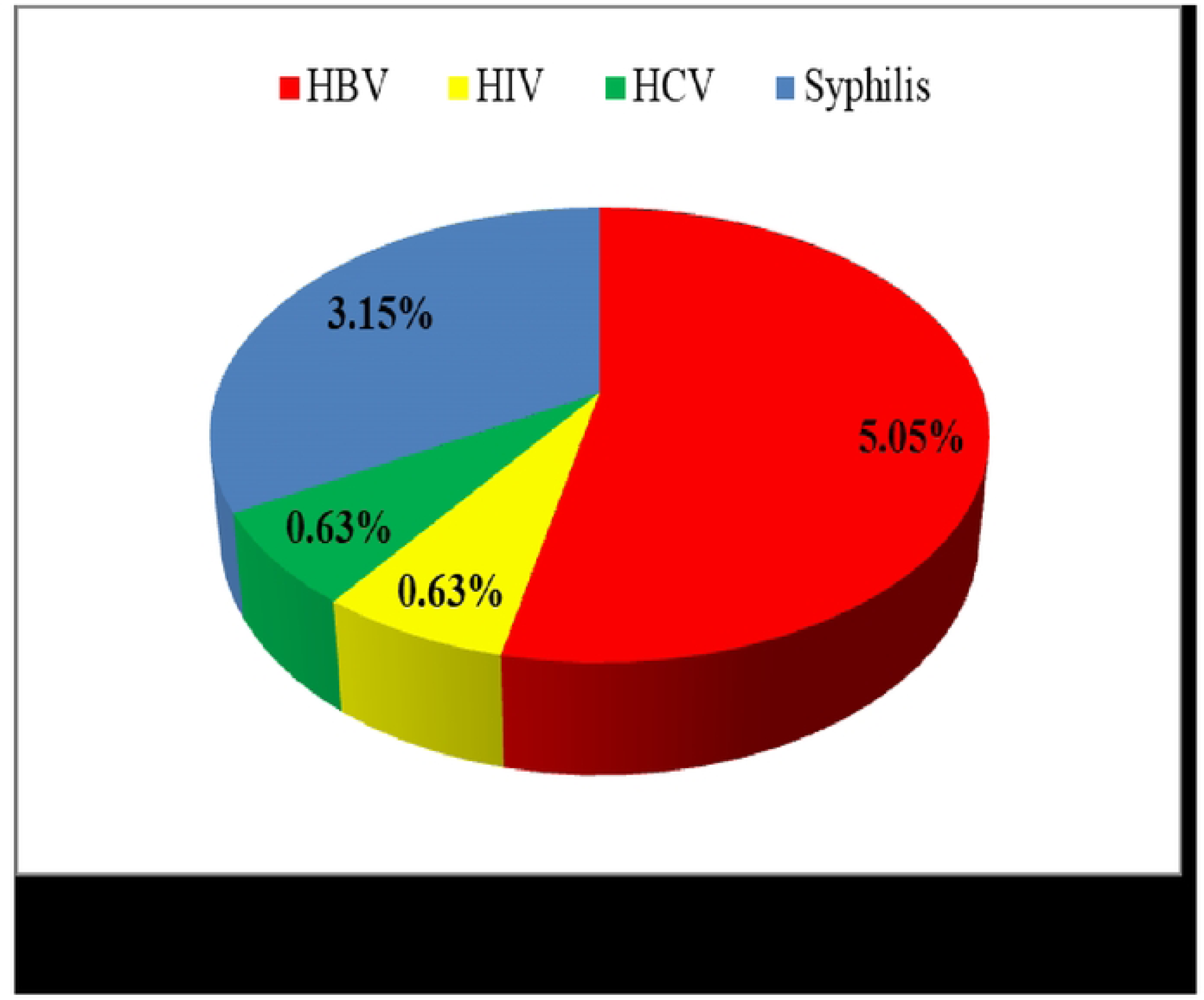

### Factors associated with transfusion transmittable infections

A binary logistic regression analysis was used to examine the associated factors of transfusion-transmissible infections. Variables with a p-value below 0.25 at bivariable logistic regression analysis were entered into the multivariable logistic regression model. Accordingly, in the crude analysis, older age, participants without formal education, hearing information about transfusion transmissible infections, knowing how transfusion transmissible infections transmit, first-time donation, receiving blood products, having sex, unprotected sexual intercourse with more partners, and history of STDs were significantly associated with transfusion transmissible infections.

Then multivariable logistic regression analysis was used to identify independent predictors for transfusion-transmitted infections and to declare statistical significance at a p-value less than 0.05. Thus, unprotected sexual intercourse with more partners is 4.77 times (AOR = 4.77; 95% CI: 1.38–16.44; P<0.013) more likely to develop a transfusion transmissible infection as compared to those who have not had unprotected sexual intercourse with more partners.

Participants with the first donation were statistically significant predictors of transfusion transmissible infections, in which the odds of having one transfusion transmissible infection among study participants were increased by a factor of 2.85 (AOR = 2.85; 95%CI: 1.16–6.99; P<0.022) as compared to those who donated repeatedly. The odds of having one transfusion transmissible infection among participants with no formal education were 4.84 times (AOR = 4.84; 95% CI: 1.09–21.46; P<0.038) higher than those of persons who learned secondary and above. (**Table 3**)

**Table 3.**
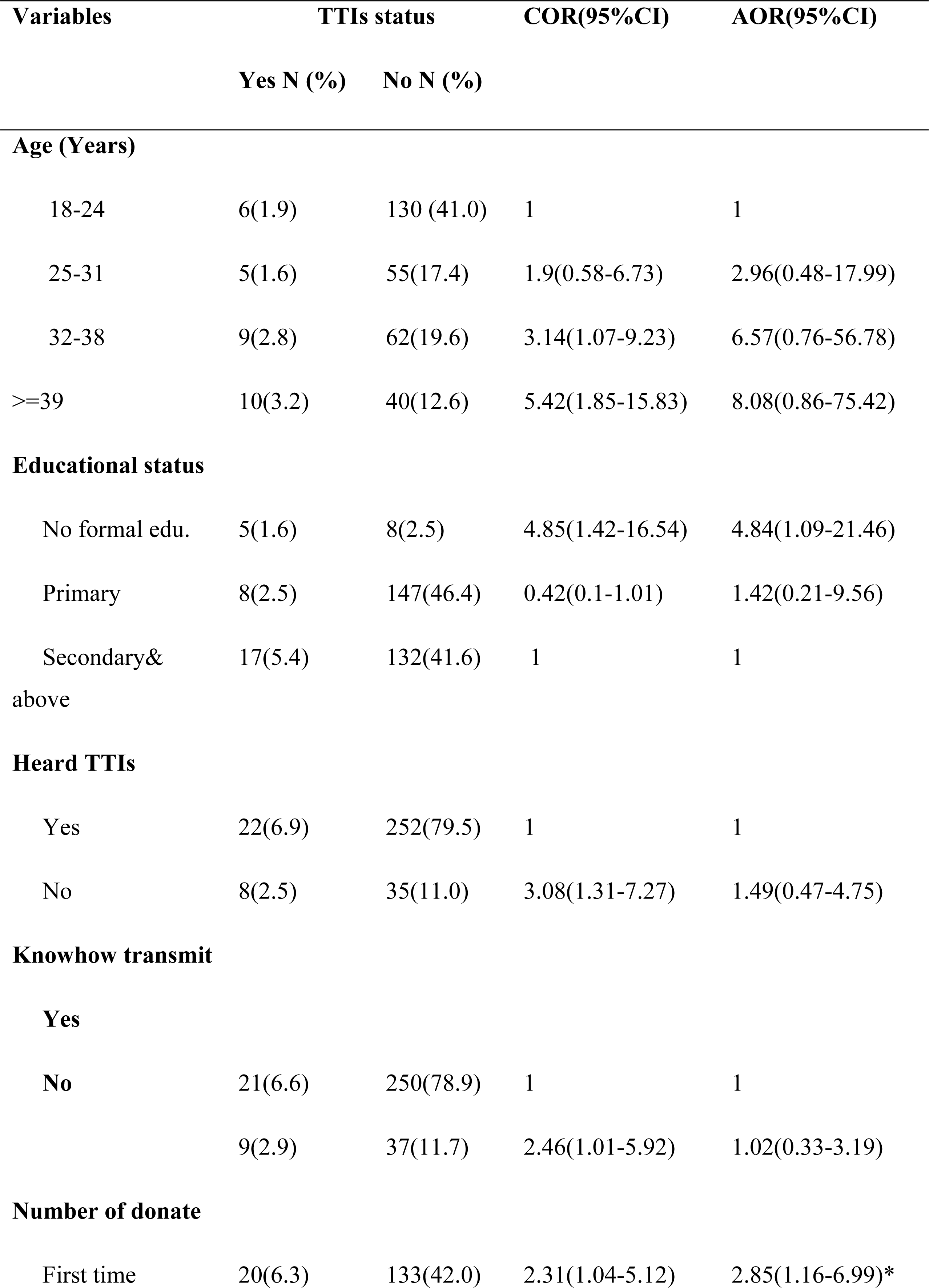
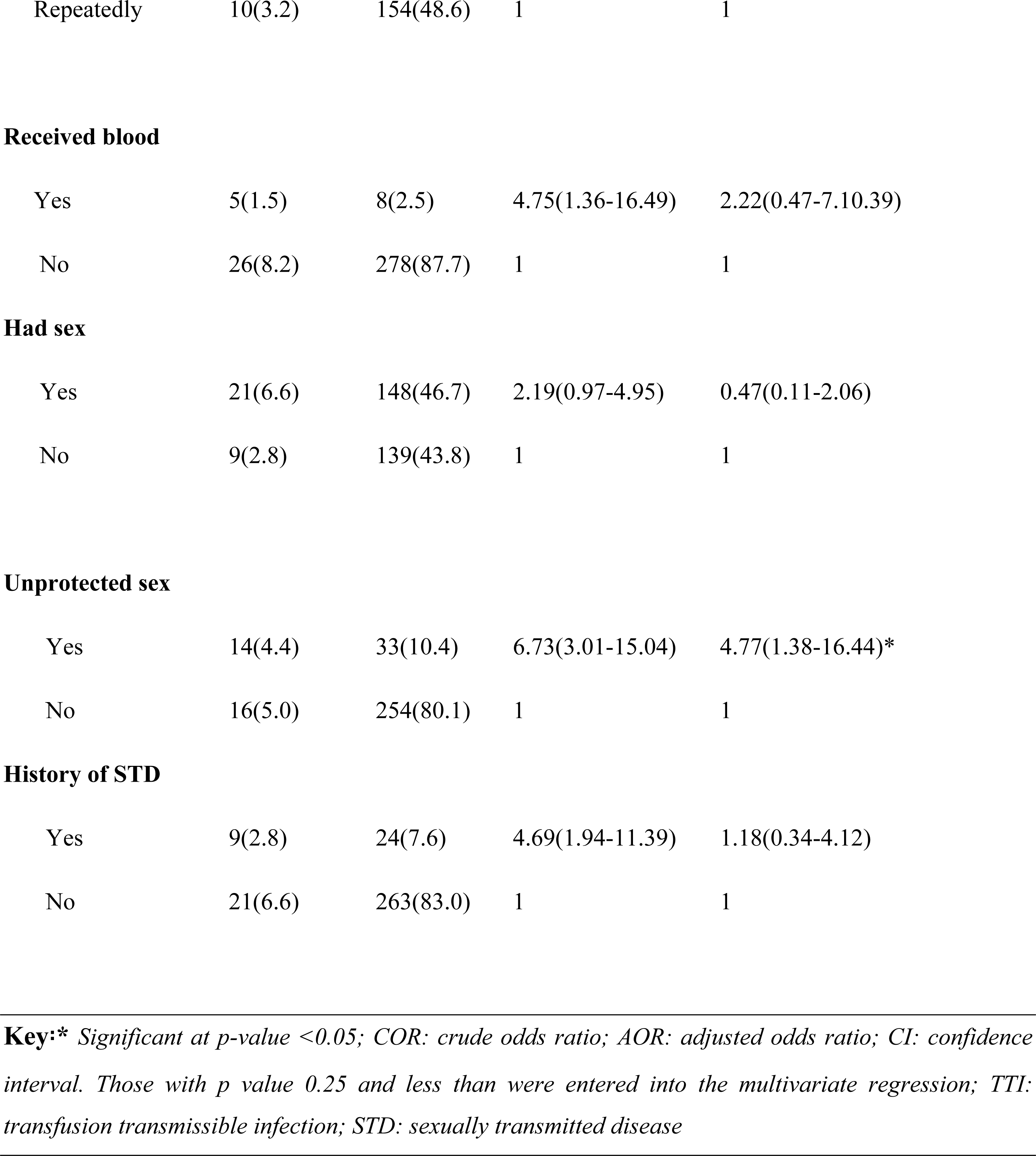
Variables associated with TTIs by both crude and adjusted odds ratio among voluntary blood donors at Woliso Blood Bank of South West Shewa, Oromia Region, Ethiopia, 2021.

| Variables | TTIs status |  | COR(95%CI) | AOR(95%CI) |
| --- | --- | --- | --- | --- |
|  | Yes N (%) | No N (%) |  |  |
| Age (Years) |  |  |  |  |
| 18-24 | 6(1.9) | 130 (41.0) | 1 | 1 |
| 25-31 | 5(1.6) | 55(17.4) | 1.9(0.58-6.73) | 2.96(0.48-17.99) |
| 32-38 | 9(2.8) | 62(19.6) | 3.14(1.07-9.23) | 6.57(0.76-56.78) |
| >=39 | 10(3.2) | 40(12.6) | 5.42(1.85-15.83) | 8.08(0.86-75.42) |
| Educational status |  |  |  |  |
| No formal edu. | 5(1.6) | 8(2.5) | 4.85(1.42-16.54) | 4.84(1.09-21.46) |
| Primary | 8(2.5) | 147(46.4) | 0.42(0.1-1.01) | 1.42(0.21-9.56) |
| Secondary&<br>above | 17(5.4) | 132(41.6) | 1 | 1 |
| Heard TTIs |  |  |  |  |
| Yes | 22(6.9) | 252(79.5) | 1 | 1 |
| No | 8(2.5) | 35(11.0) | 3.08(1.31-7.27) | 1.49(0.47-4.75) |
| Knowhow transmit |  |  |  |  |
| Yes |  |  |  |  |
| No | 21(6.6) | 250(78.9) | 1 | 1 |
|  | 9(2.9) | 37(11.7) | 2.46(1.01-5.92) | 1.02(0.33-3.19) |
| Number of donate |  |  |  |  |
| First time | 20(6.3) | 133(42.0) | 2.31(1.04-5.12) | 2.85(1.16-6.99)* |
| Repeatedly | 10(3.2) | 154(48.6) | 1 | 1 |
| Received blood |  |  |  |  |
| Yes | 5(1.5) | 8(2.5) | 4.75(1.36-16.49) | 2.22(0.47-7.10.39) |
| No | 26(8.2) | 278(87.7) | 1 | 1 |
| Had sex |  |  |  |  |
| Yes | 21(6.6) | 148(46.7) | 2.19(0.97-4.95) | 0.47(0.11-2.06) |
| No | 9(2.8) | 139(43.8) | 1 | 1 |
| Unprotected sex |  |  |  |  |
| Yes | 14(4.4) | 33(10.4) | 6.73(3.01-15.04) | 4.77(1.38-16.44)* |
| No | 16(5.0) | 254(80.1) | 1 | 1 |
| History of STD |  |  |  |  |
| Yes | 9(2.8) | 24(7.6) | 4.69(1.94-11.39) | 1.18(0.34-4.12) |
| No | 21(6.6) | 263(83.0) | 1 | 1 |

**Key:**\* Significant at $p$ -value $<0.05$ ; COR: crude odds ratio; AOR: adjusted odds ratio; CI: confidence interval. Those with $p$ value 0.25 and less than were entered into the multivariate regression; TTI: transfusion transmissible infection; STD: sexually transmitted disease

## Discussion

In this study, the overall sero-prevalence of transfusion-transmitted infections was 9.5% (95% CI: 6.3–12.9). It was similar to the study conducted at Yemen’s national blood transfusion and research center in Sana’a (8.8% (95% CI: 5.79–11.81) (16), the study in Tanzania (10.1% (95% CI: 9.92–10.28) (17), the Sidama region of Ethiopia (7.29% (95% CI: 4.69–9.89) (12) and Eastern Ethiopia (12.4% (95% CI: 9.5–15.3) (20). However, it was higher than another study conducted in Northern Ethiopia, which was 6% (95% CI: 5.75–6.25) (21) and and Harar blood bank of Eastern Ethiopia, 6.6% (95% CI: 6.14–7.06) (22) and lower than reports from Kenya, 14.1% (95% CI: 11.3–16.9) (18). These variations might be due to differences in the study population, study setting, and sample size they used to estimate the prevalence.

The sero-prevalence of HBV in this study was 5.05% (95% CI: 2.65, 7.45). This finding was similar to the study conducted in Tanzania (5.1% (95% CI: 4.97, 5.23) (17), the Regional Blood Transfusion Center Nakuru and Tenwek Mission Hospital of Kenya (6.0% (95% CI: 4.1, 7.9) (18), Eastern Ethiopia (6.6% (95% CI: 4.42, 8.78) (20), and the Sidama Region of Ethiopia (4.2% (95% CI: 2.2, 6.2) (12). However, this finding was higher than the previous systematic review reports from Pakistan 2.04% (95% CI: 0.81, 4.22) (15), Yemen’s national blood transfusion and research center in Sana’a 2.5% (95% CI: 0.84, 4.16) (16), and Harar blood bank of Eastern Ethiopia 4.4% (95% CI: 4.02, 4.78) (22), and lower than the study result from Bahir Dar district blood bank of Northwest Ethiopia 6.0% (95% CI: 5.75, 6.25) (21). The possible explanation for these variations could be due to behavioral characteristics such as unprotected sex, tooth extraction, and tattoos, which were high in these study participants. Also, HBV has a high prevalence in the general population, which arises from the high infectivity potential of the virus.

In this study, the sero-prevalence of syphilis was 3.15% (95% CI: 1.25, 5.05). This result was similar to studies conducted in Eastern Ethiopia (3.4%; 95% CI: 1.8–5.0) (20). However, it was higher as compared with other studies conducted in Kenya 2.0% (95% CI: 0.87, 3.13) (18), Sidama Region of Ethiopia 0.8% (95% CI: 0.0, 1.6) (12), Harar blood bank of Eastern Ethiopia 1.1% (95% CI: 0.91, 1.29) (22), Northern Ethiopia 1.2% (95% CI: 1.10, 1.30) (21) and Yemen’s national blood transfusion and research center in Sana’a 1.2% (95% CI: 0.00, 2.4) (16). The lower rate of sero-prevalence of syphilis in most studies might be due to the conceivable fact that syphilis is less often transmitted by blood. The duration of the study and the cultural and behavioral characteristics of the study participant might be some of the possible explanations.

The voluntary blood donors sero-prevalence of HIV in this study was 0.63%. This finding was similar to a study conducted in the Harar blood bank of Eastern Ethiopia (0.6% (95% CI: 0.46, 0.74) (22), the Sidama Region of Ethiopia (1.6% (95% CI: 0.4, 2.8) (12) and Eastern Ethiopia (1.4% (95% CI: 0.37, 2.43) (20). But it was lower than reports from Kenya (9.0% (95% CI: 6.7, 11.3) (18), and higher than study results from Yemen’s national blood transfusion and research center in Sana’a (0.3% (95% CI: 0.0, 0.6) (16) and Northern Ethiopia (0.5% (95% CI: 0.43, 0.57) (21). These discrepancies might be due to the behavioral characteristics of the study participants with different study areas and sample sizes.

In this study, the sero-prevalence of HCV was 0.63%. This finding was similar to the study conducted in Yemen’s national blood transfusion and research center in Sana’a: 1.2% (95% CI: 0.0, 2.4) (16), in eastern Ethiopia: 1.0% (95% CI: 0.13, 1.87) (20), in northern Ethiopia: 0.6% (95% CI: 0.52, 0.68) (21), and in the Sidama Region of Ethiopia: 0.5% (95% CI: 0.0, 1.0) (12). However, it was lower than reports from Kenya (8.0% (95% CI: 5.82, 10.18) (18), and the Harar blood bank of Eastern Ethiopia (0.8% (95% CI: 0.64, 0.96) (22). The possible reason might be due to the behavioral characteristics of the study participants with different study areas and sample sizes.

In this study, the odds of developing one of the TTIs were 4.85 [AOR = 4.85; 95% CI: 1.42, 16.54] times higher among voluntary blood donors with no formal education compared to those learned secondary and above. This finding is comparable with the study done in Eastern Ethiopia (20). The sero-prevalence of TTIs in this study was found to decrease with an increasing level of education. This might be attributed to the fact that as the level of education increases, there is a high probability of being aware of preventive measures against TTIs.

The study also showed that the odds of developing one of the TTIs were 4.77 [AOR = 4.77; 95% CI: 1.38, 16.44] among voluntary blood donors who had unprotected sexual intercourse compared to those who did not practice unprotected sex. This corresponds with research conducted in Eastern Ethiopia, Kenya, and South Gonder of Ethiopia (20) (25) and (26) respectively. The possible explanation could be that sexual activity with multiple partners is the key mode of acquiring sexually transmitted infections.

The study also showed that the odds of developing one of the TTIs were 2.85 [AOR = 2.85; 95% CI: 1.16, 6.99] times higher among individuals who donated for the first time compared to those who had donated repeatedly. This finding was supported by the findings of a cross-sectional study conducted in the South Gondar District Blood Bank, Northwest Ethiopia (26). The reason might be due to obtaining awareness about TTIs from post-blood donation counseling while participating repeatedly in such activities.

## Conclusion

The prevalence of at least one transfusion-transmissible infection in this study was relatively high as compared to studies conducted earlier in Ethiopia. Also, the study identified determinants of TTIs among voluntary blood donors based on socio-demographic, clinical, and behavioral characteristics. Finally, educational level, number of donations, and unprotected sexual intercourse with more partners were found to be independent predictors for one of the TTIs.

### Recommendation

Special attention need to be given to transfusion transmissible infections particularly, HBV and syphilis infections in Southwest Shewa of Oromia Region, Ethiopia. National blood bank should work in collaboration with different stakeholders and all others district blood banks in strengthening a screening plan and post donation counselling strategies for monitoring the implementation at all levels to reduce TTIs. Further prospective studies should be conducted rigorously in order to identify the cause and effect relationship of TTIs with its contributing factors. Additionally, each of the blood banks and Regional health bureaus in the study area should mobilize community for increasing repeated voluntary donors and enhance health promotion about prevention and control transmission transmissible infections by considering the identified associated factors.

### Strengths and limitations of the study

This study was intended to assess the prevalence of transfusion-transmissible infections with highly specific and sensitive laboratory methods. The methodological parts need particular attention because this study was limited by its cross-sectional study design, in which the temporal relationship between risk factors and the outcome could not be determined due to both being examined at the same time.

## Data Availability

All relevant data are within the manuscript and its Supporting Information files.

## Abbreviations

CI: Confidence Interval;
ELISA: Enzyme Linked Immune Sorbent Assay;
ETB: Ethiopian Birr;
EU: European Union;
FMOH: Federal Ministry of Health;
HBV: Hepatitis B Virus;
HBsAg: Hepatitis B Surface Antigen;
HCV: Hepatitis C Virus;
HIV: Human Immunodeficiency Virus;
NBB: National Blood Bank;
STD: Sexually Transmitted Disease;
TTIs: Transfusion Transmissible Infections;
WHO: World Health Organization

## Data availability

The data used to support the findings of this study are available from the corresponding author upon request.

## Contributions of author’s

All authors made a significant contribution to the work reported, whether that is in the conception, study design, execution, acquisition of data, analysis, and interpretation, or in all these areas; took part in drafting, revising, or critically reviewing the article; gave final approval of the version to be published; have agreed on the journal to which the article has been submitted; and agree to be accountable for all aspects of the work.

## Acknowledgments

We would like to express our gratitude to the Ambo University College of Medicine and Health Sciences Department of Public Health for giving us the opportunity to prepare this research. Finally, we would like to express our appreciation to the study participants for their willingness to provide the required information, data collectors, supervisors, and Woliso Blood Bank staff.

